# Perceptions of Open Science in the Editorial and Peer Review Process: A Cross-Sectional Survey of Traditional, Complementary, and Integrative Medicine Journal Editors

**DOI:** 10.64898/2026.06.30.26356457

**Authors:** Jeremy Y. Ng, Daivat Bhavsar, Julian T. Lau, Neha Dhanvanthry, Daniel Fry, Ji Woo Kim, Amelia King, Jaimie Lai, Anthony Makwanda, Priscilla Olugbemiro, Jeel Patel, Insha Virani, Ella Ying, Kingsley Yong, Abdullah Zaidi, Jasmine Zouhair, Susan Arentz, Erik J. Groessl, Myeong Soo Lee, Ye-Seul Lee, Ava Lorenc, L. Susan Wieland, Holger Cramer

## Abstract

**Background:** Open science (OS) offers opportunities to address challenges in the editorial and peer review processes of traditional, complementary, and integrative medicine (TCIM) journals. This study assessed TCIM journal editors’ perceptions of OS and the perceived benefits and challenges of integrating OS into editorial and peer review processes.

**Methods:** A cross-sectional survey was distributed to editors-in-chief, associate editors, and editorial board members of 115 TCIM journals. The survey examined demographics, current use and familiarity with OS, perceived advantages and obstacles, and future perspectives on OS in academic publishing. Quantitative data were analyzed descriptively, and qualitative data were examined using thematic analysis.

**Results:** A total of 267 respondents completed the survey, with most identifying as faculty members or academic research staff (n = 201/335, 60.0%). Most respondents were familiar (n = 128/212, 60.3%) or very familiar (n = 64/212, 30.2%) with OS practices, although many had received no formal OS training (n = 94/210, 44.8%).

Respondents were most familiar with open access (n = 131/213, 61.5%) and preprints (n = 92/211, 43.6%). Among the seven OS practices examined, open access was viewed most favorably, with many considering it "very important" (n = 97/206, 47.1%) and strongly agreeing that it enhances the accessibility of research findings (n = 118/195, 60.5%).

**Conclusion:** Most respondents were familiar with OS but held varying perceptions regarding the importance, advantages, and disadvantages of different OS practices. These findings may inform the development and implementation of evidence-based practices and policies that meet the needs of the TCIM research community.

## Background

### Introduction to Open Science in Publishing

Open science (OS) represents a transformative approach in the scientific community, emphasizing transparency, accessibility, and collaboration throughout the research process [1, 2]. In the realm of academic publishing, OS practices include open access to publications, sharing of data and research materials, pre-registration of studies, and open peer review [1, 3, 4]. These practices aim to enhance the reliability and reproducibility of research, facilitate broader dissemination of knowledge, and foster a more inclusive and collaborative scientific environment [1–5]. The adoption of OS in the editorial and peer review process can address challenges such as publication bias, lack of transparency, and limited access to scientific findings, ultimately contributing to the integrity and credibility of scholarly literature [6].

### Importance in Traditional, Complementary, and Integrative Medicine

Traditional, complementary, and integrative medicine (TCIM) represents a holistic approach to health that emphasizes the integration of diverse knowledge systems, cultural practices, and evidence-based therapies that fall outside conventional medical care, such as acupuncture, herbal remedies, and meditative techniques [7]. The editorial and peer review processes in TCIM journals face unique challenges due to the interdisciplinary nature of the research, a decades old reputation in the medical field as sometimes having questionable science and lower quality evidence, diversity of different health outcomes associated with various complex interventions, and the necessity for rigorous scientific evaluation to maintain credibility [1, 6, 8, 9]. OS offers significant opportunities to address these challenges by promoting transparency, reproducibility, and access to research findings. The adoption of OS practices by TCIM journals may lead to improvements in the quality and reliability of published research, foster greater trust and collaboration within the scientific community by promoting greater accountability, and ensure that valuable insights from TCIM research are accessible to a broader audience [1, 2, 6]. OS can also help in validating and standardizing methodologies, thereby improving the overall rigor and acceptance of TCIM studies [6, 10–12].

### Gap in Knowledge

Despite the potential benefits, the implementation and acceptance of OS practices in the editorial and peer review processes within TCIM journals remain underexplored. There is a lack of empirical data on how editors perceive the role of OS, its advantages, challenges, and ethical implications in this specific context [4, 13].

Understanding these perceptions is crucial for informing the development and integration of OS practices tailored to the needs of TCIM journals and addressing any concerns that may impede their adoption [13]. This knowledge gap highlights the need for comprehensive research to evaluate the current attitudes and experiences of TCIM journal editors regarding OS.

### Relevance and Impact

Exploring the perceptions of TCIM journal editors regarding OS is both timely and significant. Various conventional clinical medicine disciplines, such as oncology and immunology, have leveraged platforms such as the Open Science Framework and clinicaltrials.gov beyond RCTs to enhance the transparency, reliability, and credibility of the research published in journals by ensuring that data and methodologies are more accessible and reproducible [10–12]. Moreover, open access to research findings may similarly accelerate the dissemination of knowledge and application of TCIM practices in clinical settings [1, 6]. Addressing ethical considerations such as transparency, data privacy, and intellectual property rights is essential for gaining the trust of editors and the broader scientific community [14–16]. Insights from this study will guide the development of OS practices that are effectively aligned with the specific needs and values of the TCIM community, ultimately contributing to the advancement and utilization of TCIM research in clinical settings. In summary, this study aims to fill the existing knowledge gap by providing a comprehensive assessment of TCIM journal editors’ perceptions on OS practices and their benefits and shortcomings of integration within the editorial and peer review process.

## Methods

### Open Science Statement

This study protocol was registered and made accessible on the Open Science Framework (OSF) and is available here: https://doi.org/10.17605/OSF.IO/2QYC6. All study materials and data, such as de-identified survey responses were also uploaded to OSF and can be found here: https://doi.org/10.17605/OSF.IO/E7QV2.

The final version of the manuscript was posted as a preprint prior to submission to a peer-reviewed journal.

### Study Design

Prior to the commencement of this project, clearance and approval from the University Hospital Tübingen Research Ethics Board (REB) was obtained (REB Number: 081/2025BO2). The design of this study was an anonymous, online cross-sectional survey targeting the editorial board members of 115 journals focused on TCIM, adapted from previous survey studies led by JYN [16, 17]. The survey questions were reviewed by subject area content experts (JYN, SA, EG, MSL, YSL, AL, LSW, HC).

### Sample and Sampling Method

The inclusion criteria for this study sample are editors-in-chief, managing editors, associate/executive editors, and other editorial board members of TCIM journals that are directly involved in the peer review and/or editing processes of submitted manuscripts. TCIM journals were identified following the search criteria detailed in **Table 1**, with around 115 TCIM journals included (**Appendix 1**, on OSF at https://osf.io/e7qv2/files/ujcp8. For each journal, the research team gathered the names of all editors and editorial board members from the journal webpage. The email addresses of these editors were manually searched and extracted, independently and in duplicate. Editors who were only involved in manuscript formatting (e.g., copyeditors, technical editors), statistical editors, and non-editorial staff that do not manage TCIM content in manuscript submissions were excluded from the study. Duplicate email addresses were also removed. A total of 14 authors contributed to this task (DB, ND, DF, JK, AK, JLai, AM, PO, JP, IV, EY, KY, AZ, JZ).

**Table 1.**
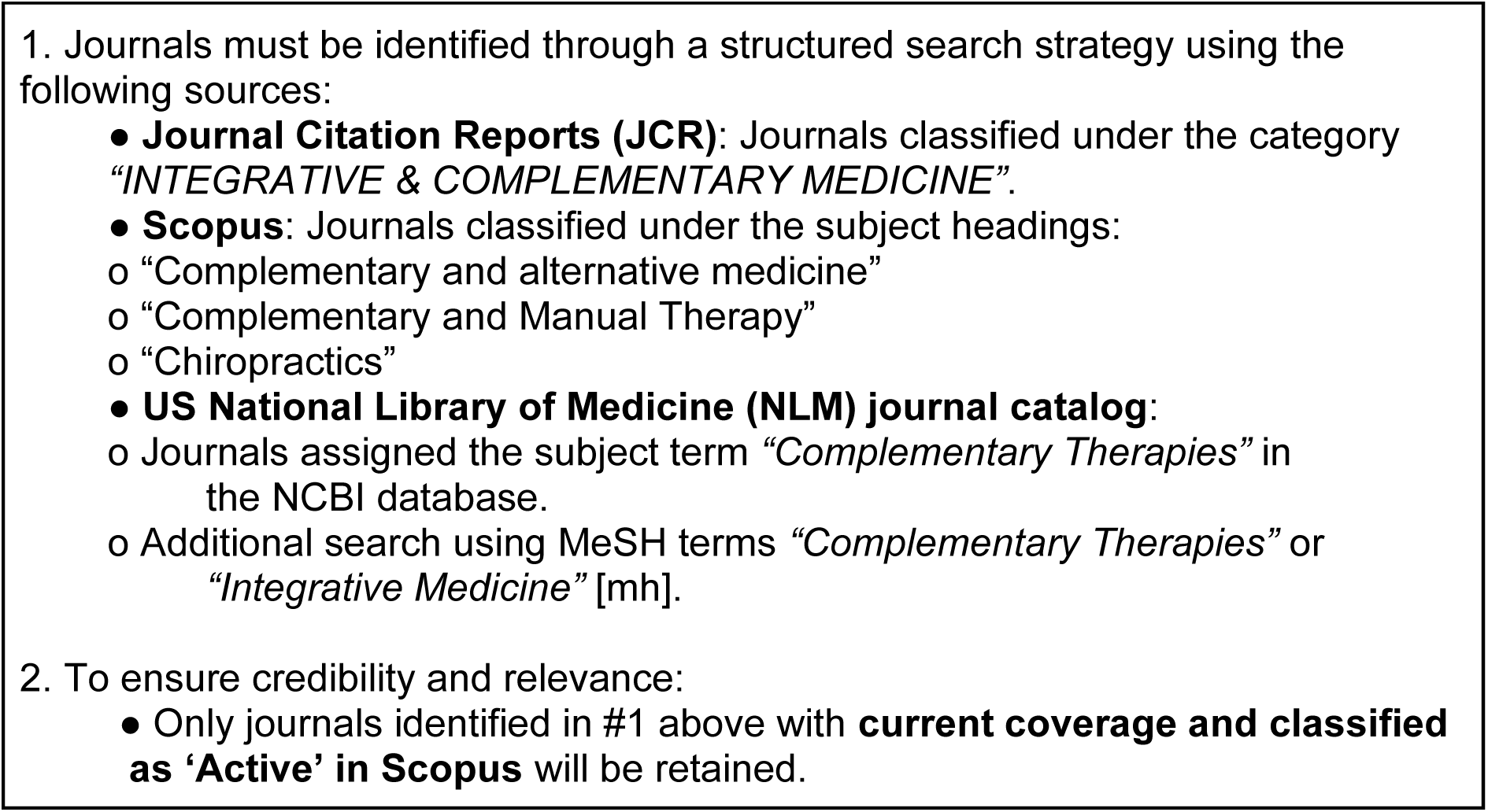
Search Criteria to Identify Traditional, Complementary, and Integrative Medicine Journals.

The survey format was closed, meaning that only invitees were able to access the survey, and they were instructed not to share the survey link with anyone else.

Emails were sent out to prospective participants via SurveyMonkey. This initial and following recruitment emails included a description of the study alongside a notice of REB approval, the objectives, and a link to access the survey, which can be found at https://osf.io/e7qv2/files/bk6se. Clicking on the survey link directed the participants to an informed consent form https://osf.io/e7qv2/files/4e29a. After the participants provided consent, participants were taken to the survey questions. Numbers of invalid and non-functional emails which bounced back were recorded by SurveyMonkey. Financial compensation or any other incentives was not provided for participation. Moreover, there was no requirement for those who were emailed a survey link to partake in this study.

### Survey Development

The survey contained a total of 44 questions, and a copy of the raw survey can be found at https://osf.io/e7qv2/files/sjgkf. The survey was designed as a structured questionnaire that begins with a screening question for eligibility and features sections on demographics and background information for potential subgroup analyses, current use and familiarity with OS in editorial tasks, perceived advantages and obstacles of OS practices, ethical considerations and potential biases, and future perspectives on OS in publishing. The survey offers descriptions for the 7 OS practices: ‘Open Access’, ‘Preprints’, ‘Open Data’, ‘Open Materials’, ‘Registration’, ‘Reporting Guidelines’, and ‘Patient and Public Involvement’. The questionnaire primarily included questions in a multiple-choice format, a matrix-style question asking participants to follow a Likert scale to describe their perceptions of the implementation of OS practices, and open-ended questions. Participants had the option to skip any questions (with the exception of the screening question) they did not wish to answer, and no personal identifying information was collected. However, once the completed survey was submitted to the study team, it was not possible to withdraw responses. The questionnaire was pilot tested with a small group of editors, and adjustments were made based on their feedback. Their feedback was incorporated as appropriate in attempts to improve the face validity of this survey.

### Data Collection

Data was collected via an online survey using SurveyMonkey (https://www.surveymonkey.com/). The initial survey email invitation was sent out on March 2nd, 2026, and remained open until April 6, 2026. Three follow-up survey reminders spaced 1 week apart (March 9, 16 and 23, 2026) were sent out in an attempt to increase response rates.

### Data Analysis

Descriptive statistics for demographic data were generated from the analysis of the reported quantitative data. Qualitative data from the open-ended responses was analysed thematically, with a data driven approach used to analyze the responses. Open-ended responses were analyzed using inductive coding and thematic analysis [18, 19]. Each response was assigned a code that captures the core meaning, which was then grouped into broader themes based on emerging patterns and commonalities. Themes were developed independently by multiple authors working in parallel to ensure reliability. An inductive approach, without reliance on pre-existing theories, guided the analysis to remain grounded in the data itself [18, 19].

The Checklist for Reporting Results of Internet E-Surveys (CHERRIES) [20] and STrengthening the Reporting of OBservational studies in Epidemiology (STROBE) were used to inform the reporting of this survey study [21].

## Results

### Demographics

Overall, 5,117 survey email invitations were sent, with 1,906 of them unopened and 684 of the emails bounced. There were 267 responses to the survey, resulting in a 6.0% (267/4433) response rate of unopened and opened email invitations, and 10.6% (267/2527) of just the opened. Raw survey data with all personal identifiers redacted (https://osf.io/e7qv2/files/5aqdr) and crosstabs (https://osf.io/e7qv2/files/y67wm) for key demographic variables (age, sex, visible minority, WHO region) have been made available on the OSF. The average time that respondents took to complete the survey was 32 minutes based on time open to start. The majority of the respondents to the survey were located in the Americas (n=86/219, 39.3%) or Europe (n=56/219, 25.6%) according to World Health Organization (WHO) World Regions. At the time of the survey, most of the respondents were TCIM journal editorial board members (n=148/219, 67.6%), followed by associate or executive directors of TCIM journals (n=71/219, 32.4%).

Many of the respondents have TCIM editorial board experience with 2-3 journals (n=90/219, 41.1%). Over half of the respondents described their career role as a faculty member or academic research staff at an academic institution (n=201/335, 60%). The research area of the manuscripts that respondents handled most frequently was clinical research (n=91/219, 41.6%). The full demographic information of the respondents can be found in **Table 2**.

**Table 2.** Characteristics of Survey Participants.

|  |  |
| --- | --- |
| **Sex** |  |
| Male | 129 (58.9%) |
| Female | 87 (39.7%) |
| Prefer not to say | 2 (0.9%) |
| Prefer to self-describe | 1 (0.5%) |
| **Age** |  |
| Under 18 | 0 |
| 18-25 | 1 (0.5%) |
| 26-35 | 5 (2.3%) |
| 36-45 | 29 (13.2%) |
| 46-55 | 65 (29.7%) |
| 56-65 | 54 (24.7%) |
| 66 or older | 63 (28.8%) |
| Prefer not to say | 2 (0.9%) |
| **Visible Minority** |  |
| Yes | 27 (12.3%) |
| No | 162 (74.0%) |
| Not sure | 22 (10.1%) |
| Prefer not to say | 8 (3.7%) |
| **World Health Organization World Region** |  |
| Africa | 7 (3.2%) |
| Americas | 86 (39.3%) |
| Eastern Mediterranean | 5 (2.3%) |
| Europe | 56 (25.6%) |
| South-East Asia | 42 (19.2%) |
| Western Pacific | 23 (10.5%) |
| **TCIM Editorial Board Experience** |  |
| 0 journals | 10 (4.6%) |
| 1 journal | 71 (32.4%) |
| 2-3 journals | 90 (41.1%) |
| 4-5 journals | 29 (13.2%) |
| More than 5 journals | 19 (8.7%) |
| **Current Role in TCIM Journal** |  |
| Editor-in-Chief (or equivalent) | 36 (16.4%) |
| Associate/Executive Editor | 71 (32.4%) |
| Managing Editor | 19 (8.7%) |
| Section Editor | 14 (6.4%) |
| Editorial Board Member | 148 (67.6%) |
| Other | 27 (12.3%) |
| **Current Career Role (Participants Could Select More Than One Role)** |  |
| Faculty member/academic research staff at a university/academic institution | 201 (60.0%) |
| Government researcher | 12 (3.6%) |
| Clinician/practitioner - conventional (MD, PharmD) | 18 (5.4%) |
| Clinician/practitioner - non-conventional (ND, DC, acupuncture, etc.) | 32 (9.6%) |
| Pharmaceutical industry/company employee | 3 (0.9%) |
| Scholarly communications employee (e.g., scholarly journals/publishing, medical writing) | 7 (2.1%) |
| Consultant (e.g., own or employed by research consulting firm) | 22 (6.6%) |
| Third sector (e.g., NGO, non-profit) | 13 (3.9%) |
| No other roles outside being an editorial board member of a TCIM journal | 2 (0.6%) |
| Other | 25 (7.5%) |
| **Primary Research Area of Handling** |  |
| Clinical research | 91 (41.6%) |
| Preclinical research – in vivo | 24 (11.0%) |
| Preclinical research – in vitro | 30 (13.7%) |
| Health systems research | 5 (2.3%) |
| Health services research | 13 (5.9%) |
| Methods research | 12 (5.5%) |
| Epidemiological research | 7 (3.2%) |
| Other | 37 (16.9%) |

### Open Science Practices and Training

In general, respondents were either “familiar” (n=128/212, 60.3%) or “very familiar” (n=64/212, 30.2%) with the concept of OS in scholarly publishing. As for the specific open practices, many respondents were “very familiar” with open access (n=131/213, 61.5%) and preprints (n=92/211, 43.6%), while respondents were “familiar” with open data (n=98/211, 46.5%) and open materials (n=79/210, 37.6%) (**Figure 1**). Many respondents were also “very familiar” with registration (n=71/208, 34.1%) and reporting guidelines (n=80/211, 37.9%), while they were “familiar” with patient and public involvement (n=60/208, 28.9%).

**Figure 1.**
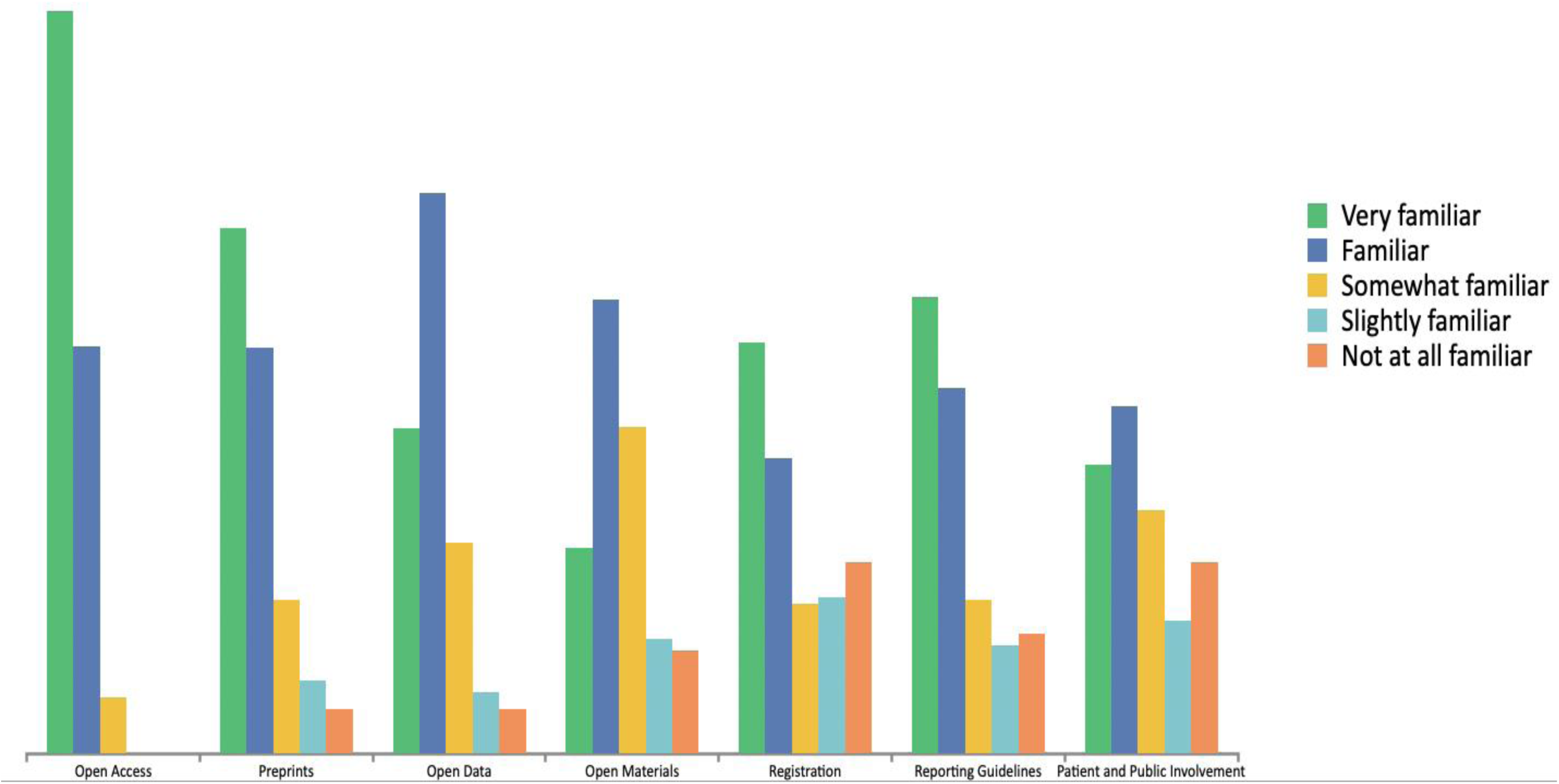
Familiarity of Respondents with Open Science Practices.

When respondents were asked about training received for these OS practices, most stated they “did not” have any formal training with respect to OS (n=94/210, 44.76%), followed closely by respondents selecting they “do” receive open science training in their primary employment (e.g. university, government) while performing peer review or editing (n=88/210, 41.9%) (**Figure 2**). Very few respondents have received training on OS through formal coursework or workshops (n=26/210, 12.4%). Many respondents indicated there was no training provided by the journals or publishers that respondents were affiliated with in regard to incorporating OS practices (n=87/210, 41.4%), followed by some respondents who stated there was training provided (n=67/210, 31.9%). Out of those respondents who indicated there was training provided to them, many of them noted that the majority of journals and publishers “do not” provide any training (n=27/67, 40.3%), followed closely by respondents indicating that the majority of journals “do” provide training (n=26/67, 38.8%). Additionally, the majority of the respondents who indicated that training was provided to them have completed the training (n=41/67, 61.2%). When asked about their preferred mode of delivering OS training, the vast majority of respondents preferred a website of resources (n=83/190, 43.7%), followed by an online webinar or recording (n=33/190, 17.4%). The majority of respondents indicated an in-person workshop hosted by their institution or journal as the least preferred format (n=104/190, 54.7%). Many respondents selected “not sure” as to whether their affiliated journals or publishers hold an official policy on how to appropriately incorporate OS practices (n=79/205, 38.5%).

**Figure 2.**
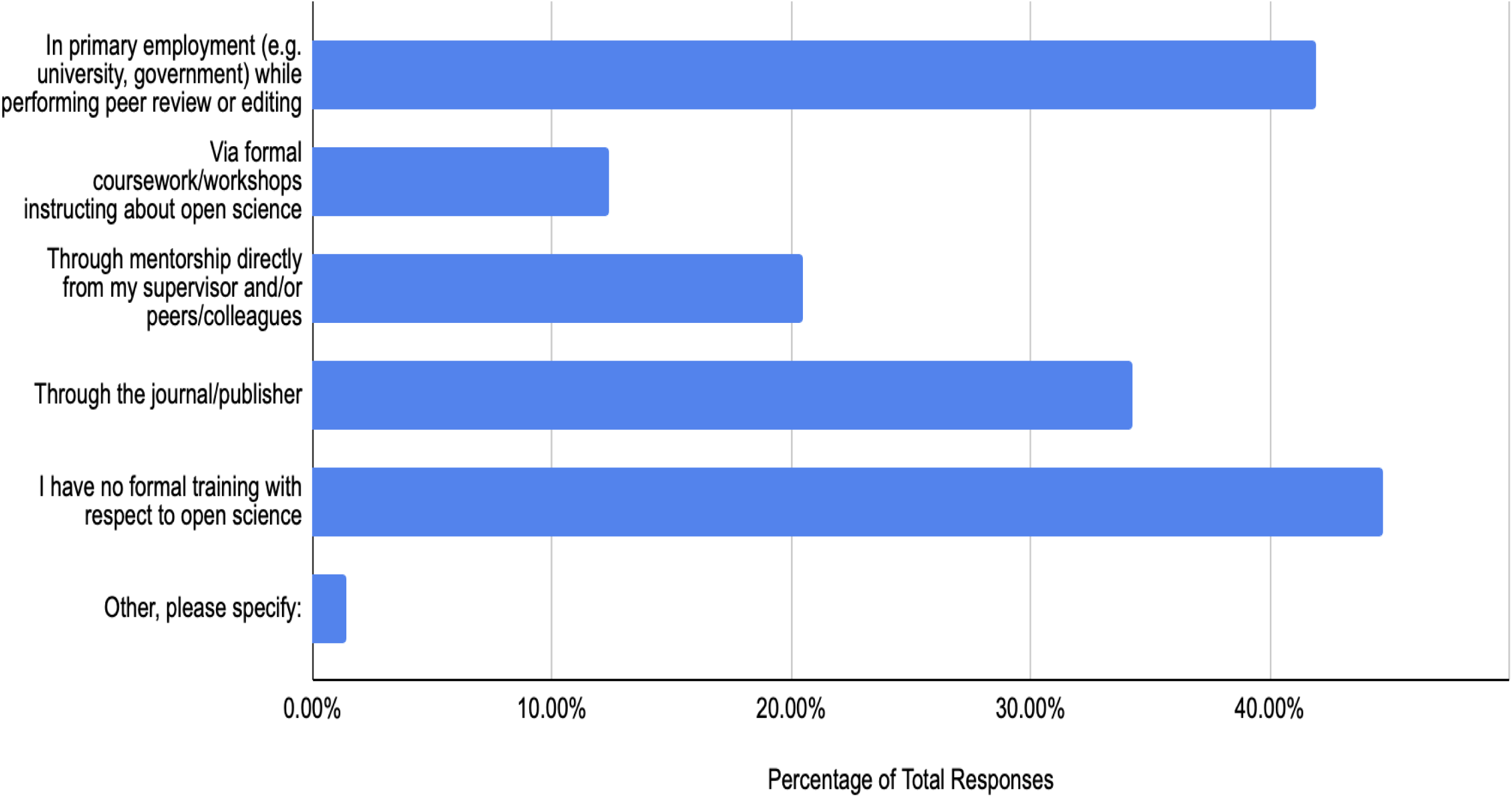
Open Science Training Received by Respondents.

### Open Access

When asked about the importance of ‘open access’, many respondents believed it to be “very important” (n=97/206, 47.1%), followed closely by “important” (n=79/206, 38.4%). The majority of respondents indicated that they “strongly agree” with open access enhancing accessibility of research findings for broader audiences (n=118/195, 60.5%) and offering the advantage of increasing transparency in research and publication (n=104/195, 53.3%) (**Figure 3**). Many respondents also indicated they “strongly agree” with open access encouraging collaboration and knowledge-sharing within the research community (n=82/195, 42.1%), accelerating the dissemination of findings within research (n=95/194, 49.0%), and building trust with readers and stakeholders by providing greater access to data and research processes (n=80/194, 41.2%). As for the disadvantages, many respondents selected “agree” that open access increases the likelihood of ethical concerns (n=62/182, 34.1%), could lead to potential misuse or misinterpretation of data by non-experts (n=59/181, 32.6%), and that it may compromise confidentiality or intellectual property of authors (n=58/181, 32.0%).

**Figure 3.**
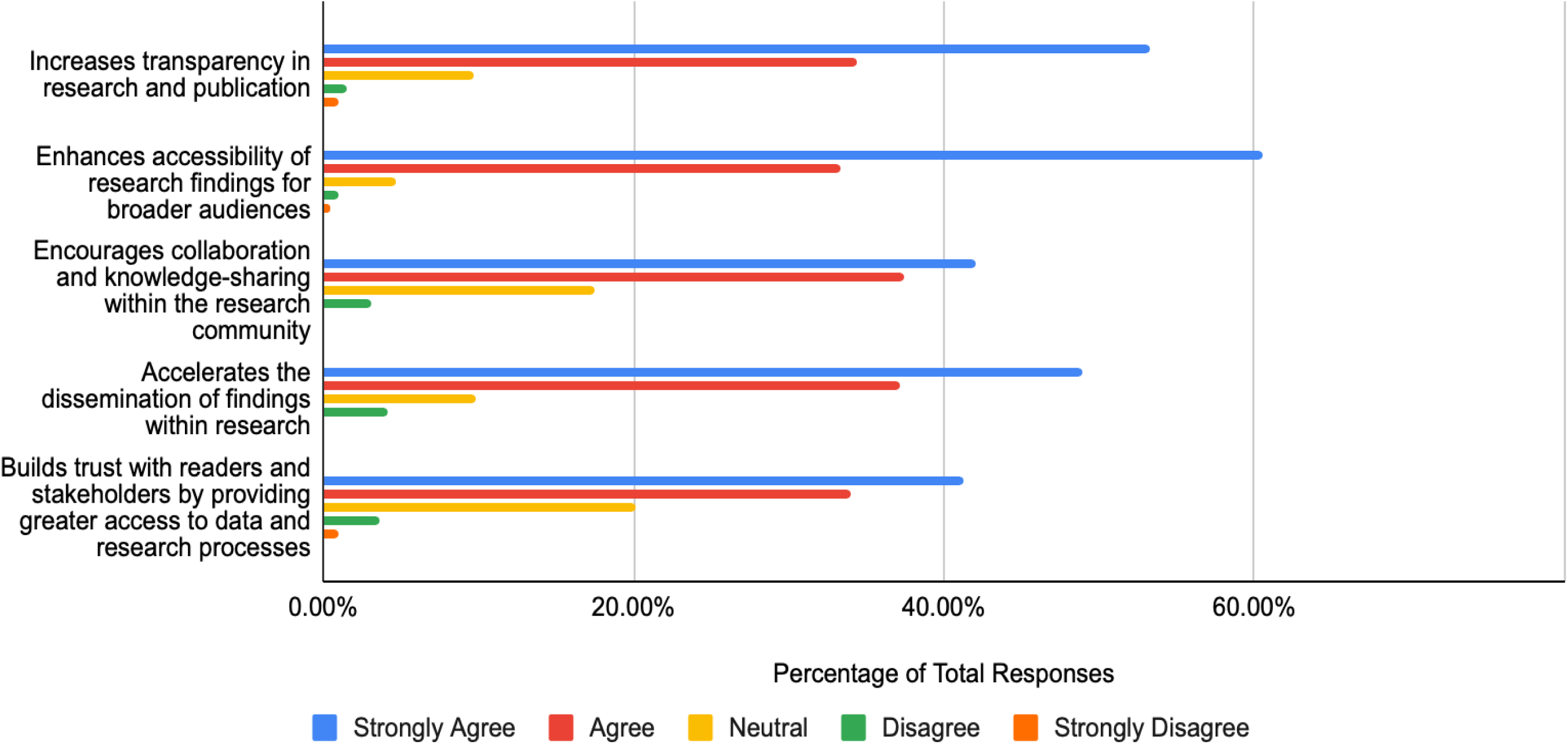
Agreement with the Following Statements Regarding ‘Open Access’ Advantages.

### Preprints

Many respondents indicated that they believe ‘preprints’ are “somewhat important” as an OS practice (n=69/205, 33.7%), followed closely by “important” (n=62/205, 30.2%). When asked about the advantages that preprints could offer, the majority of respondents selected “agree” that preprints enhance accessibility of research findings for broader audiences (n=70/193, 36.3%) and encourages collaboration and knowledge-sharing within the research community (n=74/193, 38.3%) (**Figure 4**).

**Figure 4.**
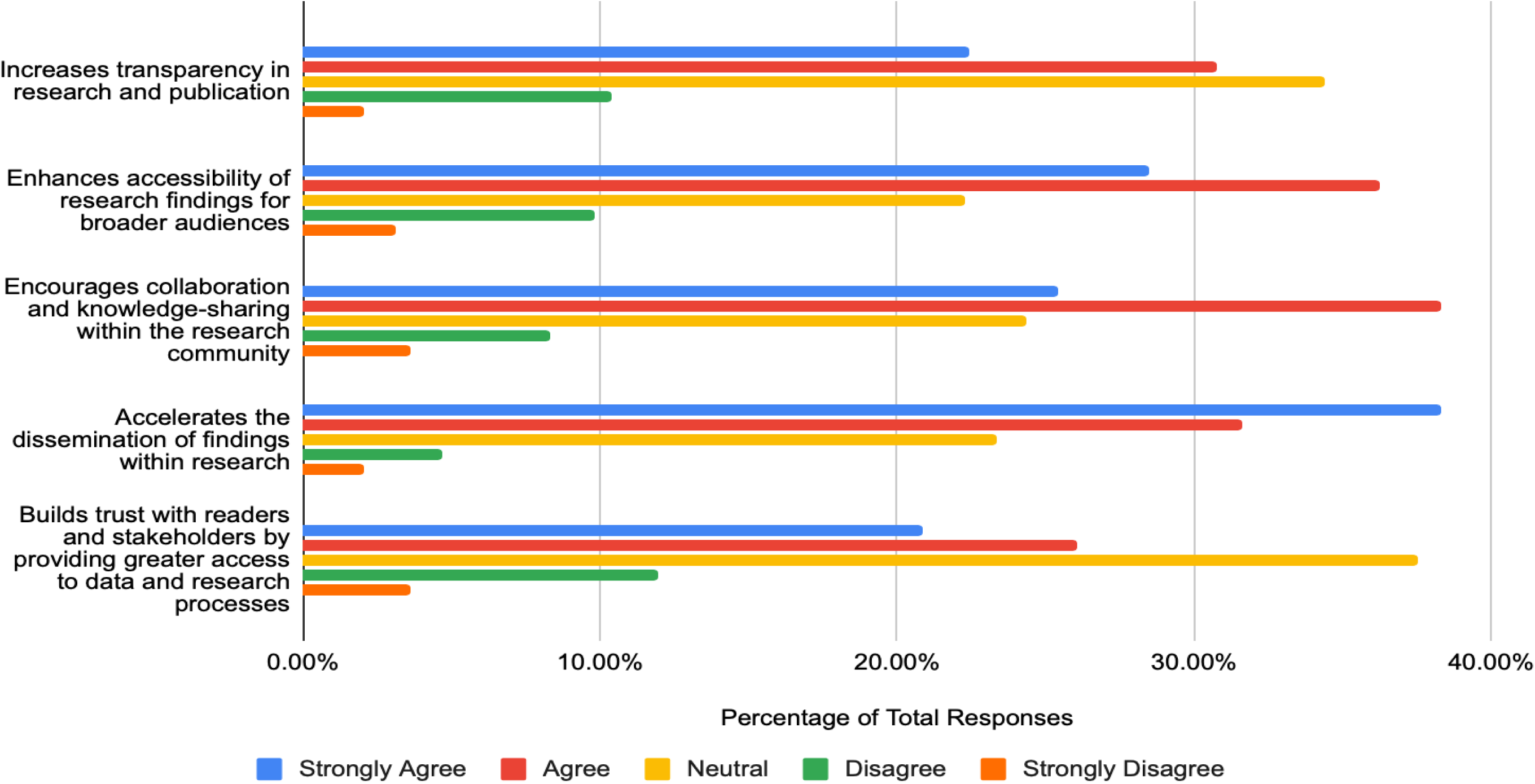
Agreement with the Following Statements Regarding ‘Preprint’ Advantages.

Additionally, many respondents selected “strongly agree” that preprints accelerate the dissemination of findings within research (n=74/193, 38.3%). As for the disadvantages, many respondents indicated that they “agree” that preprints could lead to potential misuse or misinterpretation of data by non-experts (n=65/180, 36.1%), followed by “neutral” (n=40/180, 22.2%). Respondents selected “agree” and “neutral” equally when asked if they agree that preprints pose logistical challenges in integrating with current journal policies (n=61/180, 33.9%).

### Open Data

When asked about the importance of ‘open data’ as an OS practice, many respondents selected “important” (n=85/205, 41.5%), followed by “very important” (n=68/205, 33.2%). Just under half of the respondents indicated that they “strongly agree” that an advantage of open data is that it increases transparency in research and publication (n=94/193, 48.7%) and it builds trust with readers and stakeholders by providing greater access to data and research processes (n=82/193, 42.5%) (**Figure 5**). Regarding the disadvantages of the open data, many respondents selected the option “agree” that open data could lead to potential misuse or misinterpretation of data by non-experts (n=72/179, 40.2%), may compromise confidentiality or intellectual property of authors (n=71/179, 39.7%), and increase likelihood of ethical concerns (n=70/179, 39.1%).

**Figure 5.**
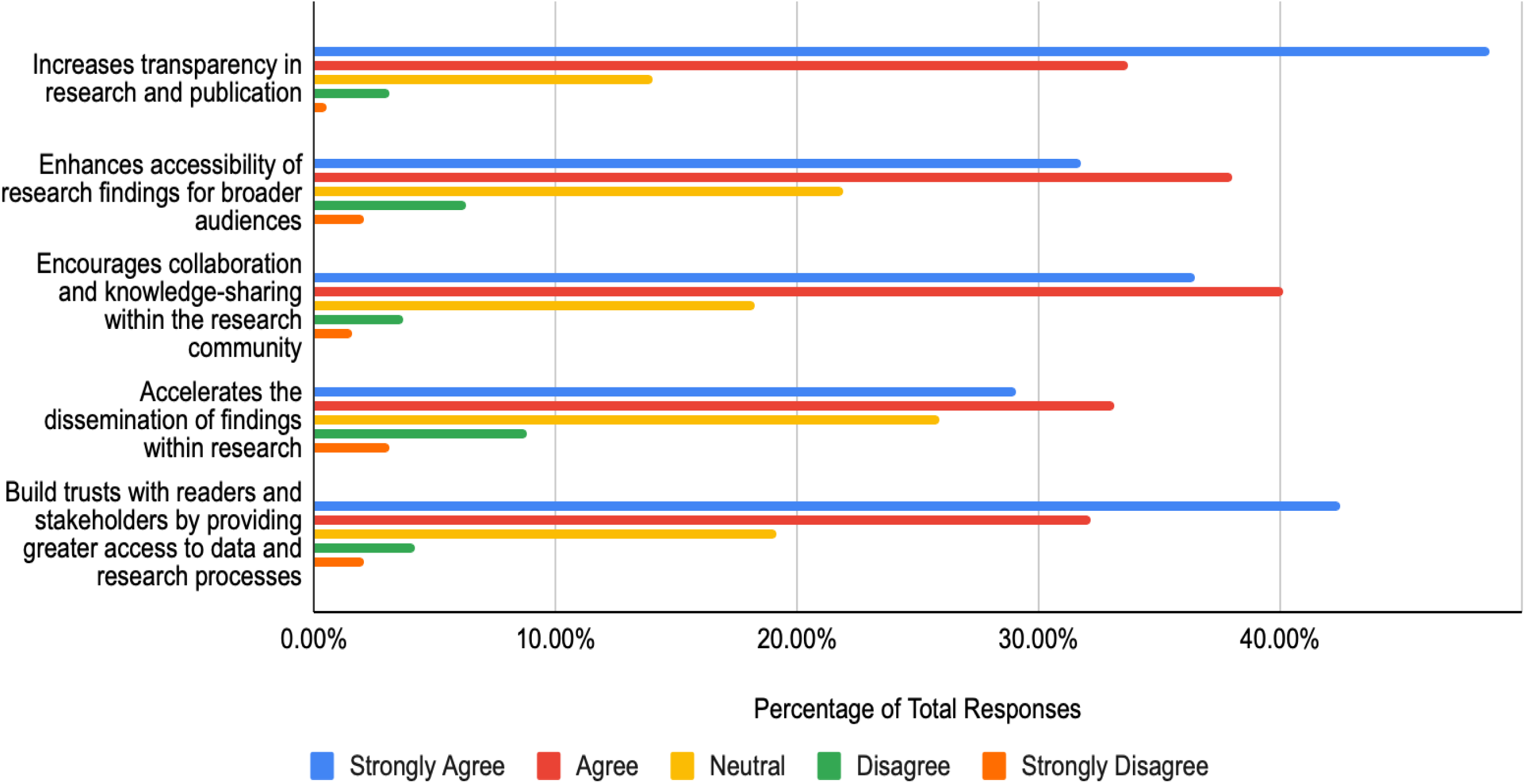
Agreement with the Following Statements Regarding ‘Open Data’ Advantages.

### Open Materials

Next, respondents were asked about the importance of ‘open materials’ as an OS practice. The majority of respondents indicated this practice as either “important” (n=79/204, 38.7%), “somewhat important” (n=54/204, 26.5%), or “very important” (n=51/204, 25.0%). When asked about the potential advantages of implementing open materials, many respondents selected “agree” that open materials encourages collaboration and knowledge-sharing within the research community (n=84/192, 43.8%) and increases transparency in research and publication (n=83/193, 43.0%). As for the disadvantages, respondents selected “neutral” to open materials posing logistical challenges in integrating with current journal policies (n=70/178, 39.3%) and to increasing the likelihood of ethical concerns (n=62/179, 34.6%). Many respondents indicated “agree” that open materials could lead to potential misuse or misinterpretation of data by non-experts (n=69/178, 38.8%) or compromise confidentiality or intellectual property of authors (n=66/179, 36.9%) (**Figure 6**).

**Figure 6.**
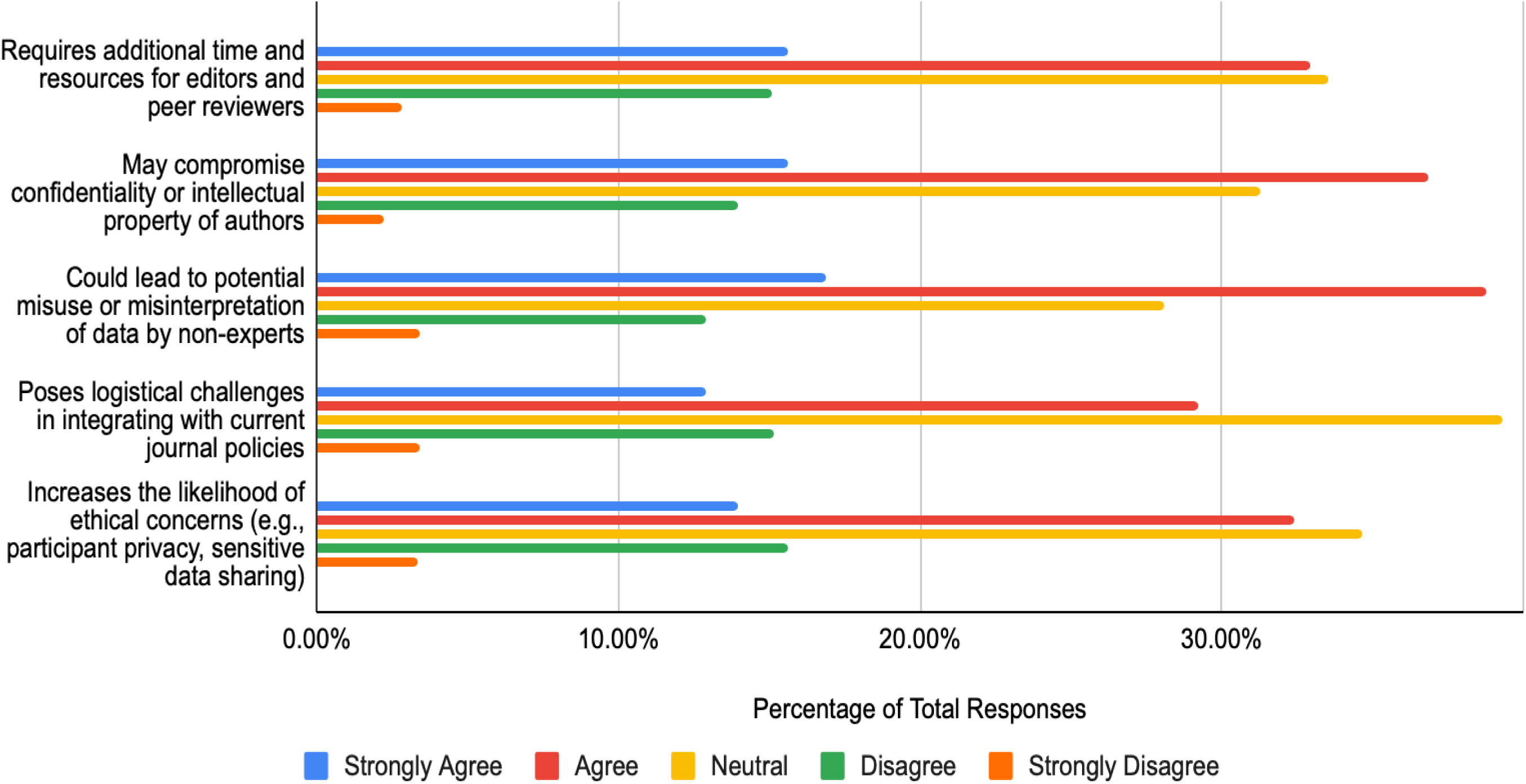
Agreement with the Following Statements Regarding ‘Open Materials’ Disadvantages.

### Registration

Many respondents believed that ‘registration’ is “very important” (n=63/203, 31.0%) as an OS practice, followed by “important” (n=61/203, 30.1%). Just below half of the respondents selected that they “strongly agree” that registration increases transparency in research and publication (n=82/192, 42.7%). Many respondents selected “agree” that registration builds trust with readers and stakeholders by providing greater access to data and research processes (n=74/190, 39.0%). When given potential disadvantages of registration, many respondents selected they were “neutral” that registration poses logistical challenges in integrating with current journal policies (n=78/178, 43.8%), may compromise confidentiality or intellectual property of authors (n=71/178, 39.9%), and increases the likelihood of ethical concerns (n=70/178, 39.3%) (**Figure 7**).

**Figure 7.**
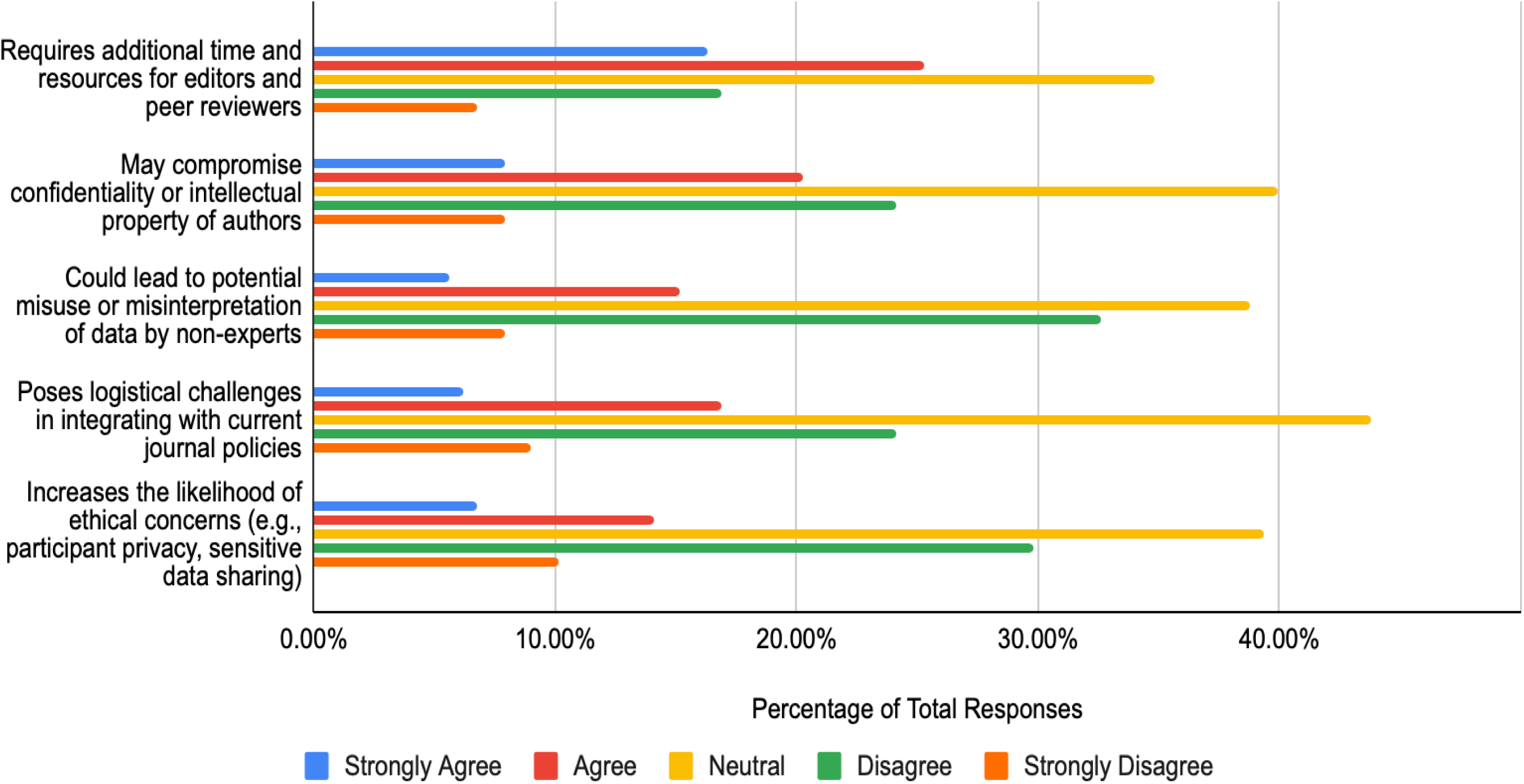
Agreement with the Following Statements Regarding ‘Registration’ Disadvantages.

### Reporting Guidelines

When asked about the importance of ‘reporting guidelines’ as an OS practice, the majority of respondents indicated that it was “important” (n=83/204, 40.7%), or “very important” (n=82/204, 40.2%). Many respondents selected that they “strongly agree” that reporting guidelines increase transparency in research and publication (n=78/191, 40.8%) and build trust with readers and stakeholders by providing greater access to data and research processes (n=72/191, 37.7%). When provided with some potential disadvantages, many respondents selected “disagree” that reporting guidelines may compromise confidentiality or intellectual property of authors (n=68/178, 38.2%), could lead to potential misuse or misinterpretation of data by non-experts (n=68/178, 38.2%), and increase the likelihood of ethical concerns (n=65/178, 36.5%) (**Figure 8**). Many respondents indicated they “agree” that reporting guidelines require additional time and resources for editors and peer reviewers (n=63/178, 35.4%).

**Figure 8.**
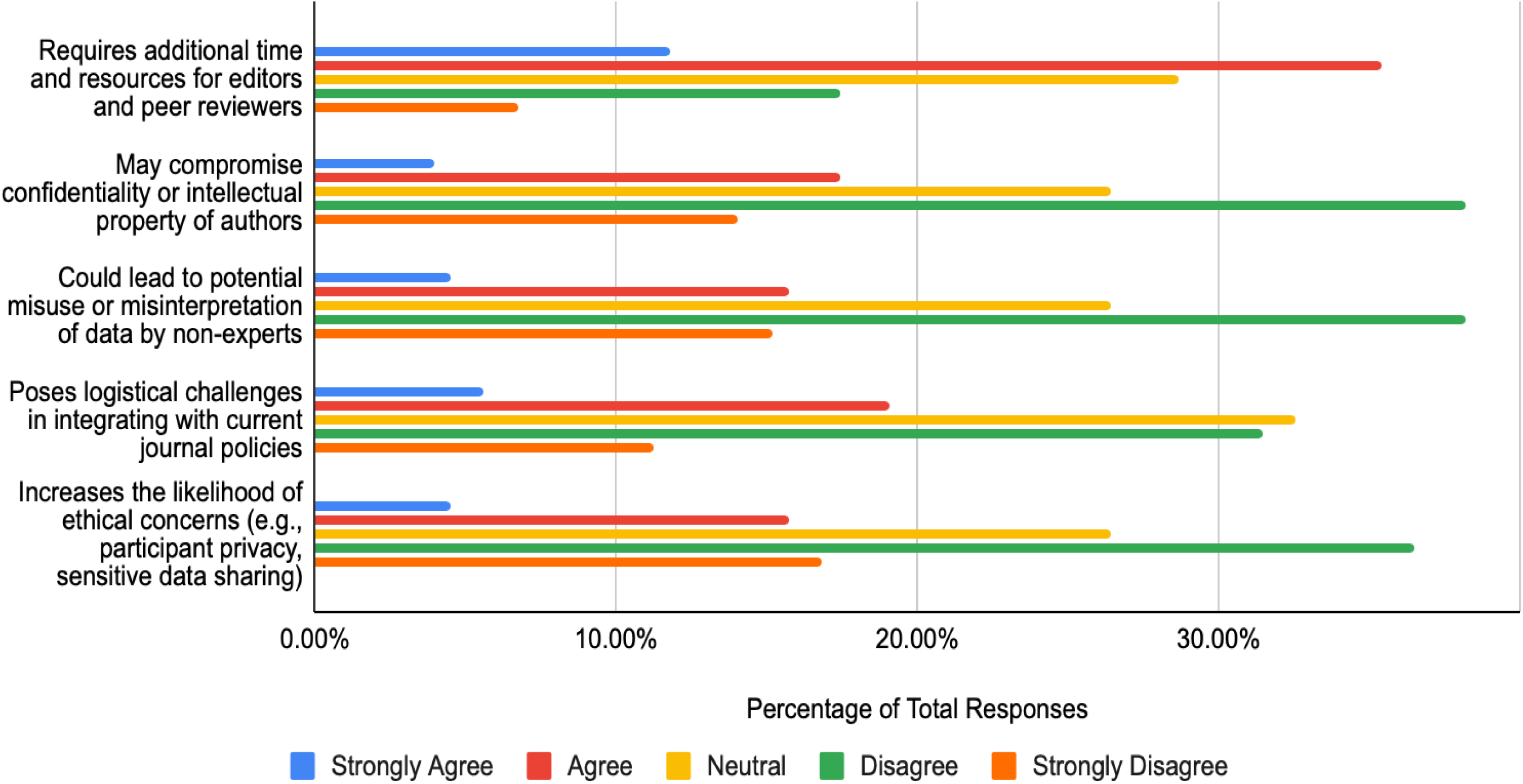
Agreement with the Following Statements Regarding ‘Reporting Guidelines’ Disadvantages.

### Patient and Public Involvement

Respondents were finally asked about the importance of ‘patient and public involvement’ as an OS practice, in which the majority of respondents selected “important” (n=68/205, 33.2%), or “somewhat important” (n=60/205, 29.3%). When asked about potential advantages, many respondents selected “neutral” that patient and public involvement accelerates the dissemination of findings within research (n=72/191, 37.7%) and encourages collaboration and knowledge-sharing within the research community (n=69/191, 36.1%). As for potential disadvantages, many respondents selected “neutral” that patient and public involvement may compromise confidentiality or intellectual property of authors (n=67/177, 37.9%), poses logistical challenges in integrating with current journal policies (n=67/177, 37.9%), and could lead to potential misuse or misinterpretation of data by non-experts (n=63/177, 35.6%) (**Figure 9**).

**Figure 9.**
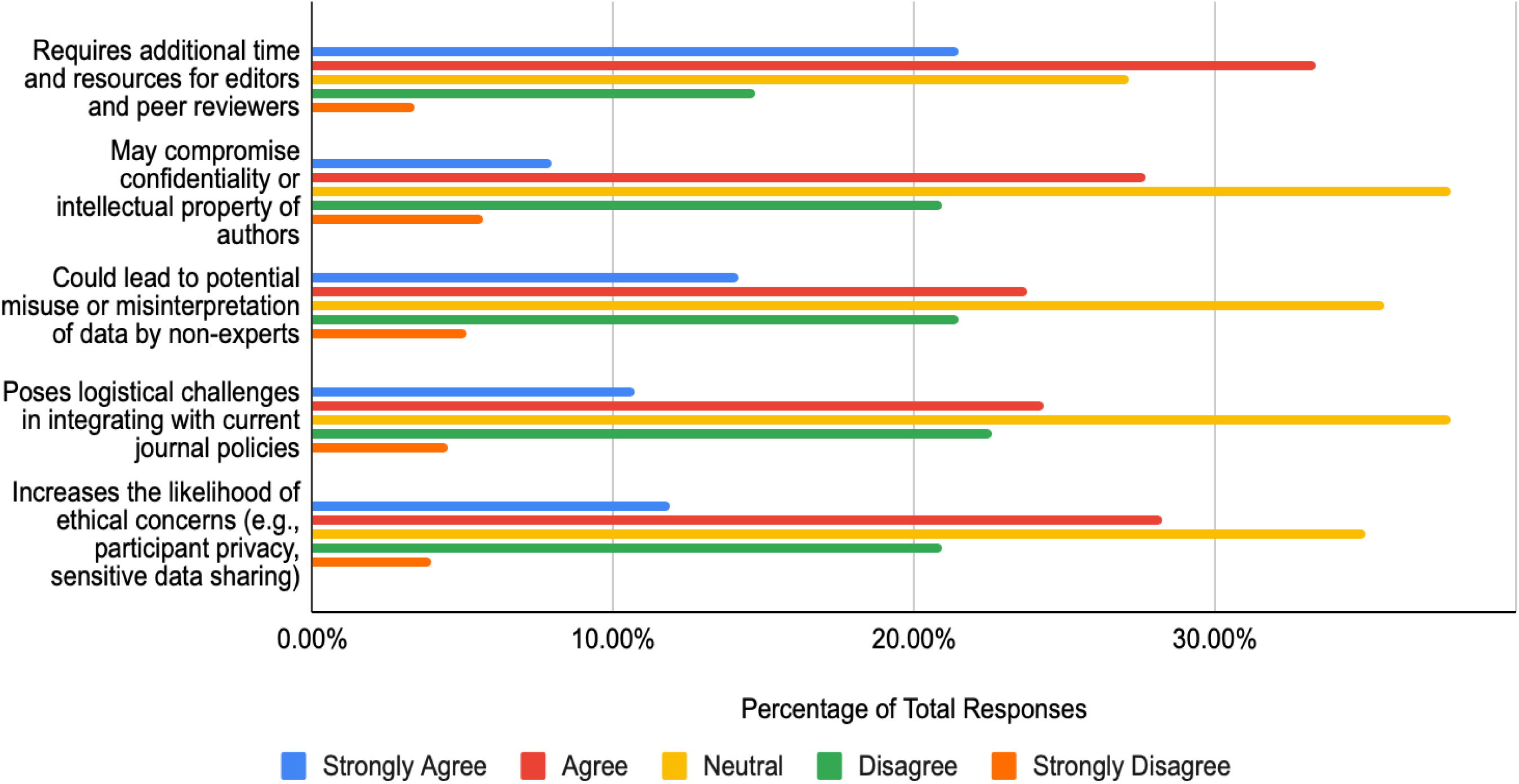
Agreement with the Following Statements Regarding ‘Patient and Public Involvement’ Disadvantages.

### Thematic Analysis and Representative Quotes

The results yielded 8 codes from 83 open-ended responses that were received. Out of these codes, 4 distinct themes were generated, each encompassing patterns identified in the dataset. Firstly, “barriers to reputable research publication” include the challenges that researchers face in publishing under the OS framework. Next, the “concerns about open science integration and TCIM on traditional STEM research and the general public” refer to responses that express concern over the integration of TCIM research with traditional STEM research and the general public through OS practices. “Concerns about the quality of outputs that come from open science practices and processes” summarized responses who believed that the research outputs through OS lacked scientific quality, leading to misinterpretations of the results. Finally, “guidelines and transparency in open science practices” reflects responses that address guideline and transparency changes in OS practices.

Definitions and representative quotes are provided in **Table 3**, while the code and thematic analysis are available at https://osf.io/e7qv2/files/8qv4z.

**Table 3.** Thematic Analysis, Definitions, and Representative Quotes.

| Themes | Definition | Representative Quotes |  |
| --- | --- | --- | --- |
| Barriers to reputable research publication | This theme refers to the challenges that researchers face in publishing under the open science framework. | “A major block to open science is financial. For publishers, there is the business concern of how to support it and the development of publication fees. For junior researchers, it puts them at a distinct cut and unfair disadvantage to getting their work out.” | “With the advent of so many predatory journals, the whole concept of open science has been abused - such as listing reviewers or having or prints before the science is all really wetted and asking huge amounts of money for "open access," e.g. prepaid distribution without real peer-review has nearly destroyed the whole system in my opinion. And then these articles are used to train AI in what "real science" reports.” |
| Concerns about open science integration and TCIM on traditional STEM research and the general public | This theme refers to the concern over the integration of TCIM research with traditional STEM research and the general public through open science practices. | “There’s still a huge gap between STEM papers, which always follow the same rules and standards, and papers in Medical Humanities coming from GLAM scholars, who follow the guidelines for Historians of science, unknown to STEM scientists, and who often work on translations as they don’t know ancient languages and have no idea of the historical background. | “Open Science always has to be taken to the public very cautiously. It is a double edged sword. In literate communities the essence of the scientific fact will be taken true to the facts while in others there may be misinterpretations.” |
| Concerns about the quality of outputs that come from open science practices and processes | This theme reflects responses who believed that the research outputs through open science lacked scientific quality, leading to misinterpretations of the results. | “We saw that pre-prints during the COVID pandemic led to the misinterpretation of data and studies - regardless of TCIM or biomedical status of a manuscript.” | “A major challenge for editors is that different clinical reviewers often have very different critiques of submitted articles, and statisticians often offer formal criticism but lack sufficient knowledge of clinical circumstances to design realistic study designs. In addition to RCTs, these clinical study results should increasingly be compared with real-world data so that guidelines also receive real-world evidence.” |
| Guidelines and transparency in open science practices | This theme addresses guideline and transparency changes in open science practices. | “Guidelines though encompassing must not cause a restriction from publication. e.g. Insisting that studies only registered in a registry, or papers only in a very specific format prescribed by the reporting guideline only will be published. Whatever work done must get published and documented in the public domain allowing the scientific community to judge its correctness, and<br>quality. Issues such as confidentiality and anonymity are seen differently in different cultures. Open access allows for greater visibility and therefore less policing is needed from journal editors or registry consultants/ editors." | “It depends on funds, manpower, time constraints, etc. Lots of revamping of journal interface may be required. It will be an additional burden but at the same time transparency can be bought into the submission process. Overall it can be a good initial step.” |

## Discussion

The primary objective of this study was to assess the perceptions of editors of TCIM journals regarding the benefits and shortcomings in integrating OS practices within the editorial and peer review process. By exploring these perceptions, the study identified perceived benefits and challenges associated with OS in academic publishing, providing valuable insights into the acceptance and potential barriers to OS implementation [1, 6, 16]. It was found that the majority of respondents are aware of OS, but have varying perceptions on the importance, advantages, and disadvantages of specific OS practices. However, respondents generally agree that there are still many barriers that need to be addressed prior to the implementation and integration of OS with the overall scientific community.

### Comparative Literature

The results of our study align with previous literature that have explored perceptions of journal editors on OS as well as the prevalent challenges and barriers to implementing OS practices in the future. In a previous cross-sectional study conducted by Melero et al. that sampled editors from all types of scientific journals, the vast majority of respondents completely agreed that open access and open data should be common practices in the peer review and editorial process [22]. This is consistent with our findings, as respondents largely felt that open access and open data were “important” or “very important”. Additionally, in interviews conducted by Melero et al., advantages of open access and open data that were consistently brought up include transparency, accessibility, validity, and reliability. In our study, respondents also felt that these were major advantages of open access and open data, with many of them indicating that they “agree” or “strongly agree” with these same advantages. In another study that surveyed editors of intervention research journals about transparency and openness in the editorial and peer review process, most of the respondents supported transparency and openness at their journals and perceived other editors in their discipline to support transparency and openness in their journals too [23]. This is consistent with our findings, as the majority of the respondents to our survey believed open data to be “important” or “very important”, and registration to also be “very important” or “important”. However, it is interesting to note that in a separate survey study that sampled editors from all types of journals, they found that there was little support for registration amongst their respondents, which is different from what we observed in our study [24]. This difference may suggest that TCIM journal editors are particularly aware of the role of a priori registration in strengthening research credibility, especially in a field where methodological quality and selective reporting remain important concerns.

Nevertheless, this interpretation should be made cautiously, as the observed differences may also reflect variations in survey samples, journal scope, disciplinary norms, or question wording across studies.

Another study that surveyed journal editors found that respondents were moderately familiar with OS, which partially aligns with the findings that respondents were “familiar” with OS in our study [13]. While the previous study sampled journal editors from all fields and types of journals, our survey only sampled TCIM journal editors, potentially accounting for the difference in the findings. With regards to training, a perspective piece observed that a lack of training and educational support was present in the TCIM community, partially due to the lack of TCIM programs at public universities in Western countries, creating a challenge in conducting TCIM research as well as engaging properly in OS practices [25]. This observation was backed up in a study that surveyed TCIM journal editors on AI usage in the editorial and peer review process, which found barriers present with lack of formal training present in the TCIM academic landscape [26]. Both observations are in line with our study, as very few respondents indicated that they have received any formal training on OS. Many respondents also indicated in our survey that the TCIM journals they are affiliated with do not offer any sort of formal training on OS.

With regards to the results of the thematic analysis, recurring themes included lack of funding and the rise in prevalence of predatory journals, both of which raise the barrier for a reputable publication and make it difficult to select a high-quality journal. Previous literature aligns with our findings, with studies stating that there are no specific funding streams for TCIM research in many countries [27,28], with student tuition providing the majority of funding at TCIM educational institutions [27]. Other studies found that journal editors agree that the emergence of predatory journals jeopardizes the credibility of open access [22].

### Strengths and Limitations

The methodological strengths of this study include its comprehensive approach to data collection and the targeted sampling method. By utilizing a list of TCIM journals and manually gathering the names and email addresses of editors, the study ensured a robust and representative sample of the target population. The structured survey, validated through pilot testing, facilitated the collection of detailed and relevant data on editors’ perceptions of OS. The survey touches upon many different prevalent OS practices, their importance, advantages, and disadvantages, providing a robust dataset that covers diverse themes. Another strength of our study lies in the diversity of respondent demographics in our survey. With strong participation from all WHO world regions and respondents from diverse career backgrounds with wide-ranging TCIM editorial board experience, our interpretations of the results are able to be better generalized to the overall target population of TCIM journal editors.

However, the study also has certain limitations, especially involving biases associated with the methodology implemented. The reliance on self-reported data may introduce response bias, as editors who are more interested in, supportive of, or opposed to OS may be more likely to participate in the survey [26]. Another example is recall bias, as respondents may not accurately remember whether they have received OS training in the past and what the training exactly consisted of, given that the survey relies on self-reporting. Non-response bias may also be observed in our results as the survey participation rate was low, possibly due to participants losing access to their email, changing their affiliation, or were on vacation, retired, or passed away. The survey was also designed, written, and administered solely in English. As a result, this may have limited or excluded potential participants who do not speak English from completing the survey and providing their views on OS integration in the editorial and peer review process.

## Conclusion

The aim of this study was to provide important insights into the perceptions of TCIM journal editors regarding the benefits and shortcomings of adopting OS practices within the editorial and peer review workflow. Respondents were provided with the opportunity to describe their training and past experiences with open science and also shared their views on the potential advantages and disadvantages of seven different open science practices. It was found that the vast majority of respondents were “very familiar” or “familiar” with open science practices, and that open access is the most widely understood open science practice. Additionally, open access was believed to be the most important as an open science practice, serving as a basis to understand what open science practices should be integrated first and how well they may be received by TCIM journal editors. These findings also help inform tailored strategies to enhance transparency, rigour, and trust in TCIM publishing. Future directions should explore the TCIM journals and editors that have already taken a shift towards the implementation of open science practices in order to further understand the benefits and challenges from the perspectives of those who have experienced the integration process of open science practices. Ultimately, the outcomes presented here in this study support the responsible integration of open science in TCIM, contributing to the advancement of both the field and the broader goals of scholarly integrity.

## List of Abbreviations

CHERRIES: Checklist for Reporting Results of Internet
E-Surveys OS: open science
OSF: Open Science Framework
REB: research ethics board
STROBE: STrengthening the Reporting of OBservational studies in Epidemiology
TCIM: traditional, complementary, and integrative medicine

## Declarations

### Ethical Approval and Consent to Participate

Prior to the commencement of this project, clearance and approval from the University Hospital Tübingen Research Ethics Board (REB) was obtained (REB Number: 081/2025BO2).

### Consent for Publication

All authors consent to this manuscript’s publication.

### Availability of Study Materials and Data

All relevant study materials and data are included in this manuscript or posted on the Open Science Framework: https://doi.org/10.17605/OSF.IO/E7QV2

### Authors’ Contribution

Jeremy Y. Ng: Conceptualization, Methodology, Investigation, Writing - Original Draft, Writing - Review & Editing, Supervision, Project administration

Daivat Bhavsar: Investigation, Writing - Original Draft, Writing - Review & Editing Julian T. Lau: Investigation, Writing - Original Draft, Writing - Review & Editing Neha Dhanvanthry: Investigation, Writing - Review & Editing

Daniel Fry: Investigation, Writing - Review & Editing Ji Woo Kim: Investigation, Writing - Review & Editing Amelia King: Investigation, Writing - Review & Editing Jaimie Lai: Investigation, Writing - Review & Editing

Anthony Makwanda: Investigation, Writing - Review & Editing Priscilla Olugbemiro: Investigation, Writing - Review & Editing Jeet Patel: Investigation, Writing - Review & Editing

Insha Virani: Investigation, Writing - Review & Editing Ella Ying: Investigation, Writing - Review & Editing Kingsley Yong: Investigation, Writing - Review & Editing Abdullah Zaidi: Investigation, Writing - Review & Editing

Jasmine Zouhair: Investigation, Writing - Review & Editing

Susan Arentz: Methodology, Investigation, Writing - Review & Editing Erik J. Groessl: Methodology, Investigation, Writing - Review & Editing Myeong Soo Lee: Methodology, Investigation, Writing - Review & Editing Ye-Seul Lee: Methodology, Investigation, Writing - Review & Editing

Ava Lorenc: Methodology, Investigation, Writing - Review & Editing

L. Susan Wieland: Methodology, Investigation, Writing - Review & Editing Holger Cramer: Methodology, Investigation, Writing - Review & Editing

## Data Availability

All relevant study materials and data are included in this manuscript or posted on the Open Science Framework.

https://doi.org/10.17605/OSF.IO/E7QV2

